# Deep Learning-Based Classification of Melanoma and Cutaneous Lesions Using NFNet Architecture: Development and Clinical Validation

**DOI:** 10.1101/2025.11.15.25340317

**Authors:** Daniel Cavadia, Marcel Castro, Margarita Oliver Llull

## Abstract

Melanoma remains the most lethal form of skin cancer, necessitating early detection for optimal patient outcomes. This study presents an advanced automated diagnostic system utilizing state-of-the-art convolutional neural networks—NFNet-L0, ResNeSt-101e, and MogaNet-XT—to classify nine types of cutaneous lesions from dermoscopic images. Leveraging a diverse dataset of 22,618 images from the International Skin Imaging Collaboration (ISIC) and Venezuelan clinical centers, the system demonstrates 96.2% accuracy in multi-class classification, outperforming conventional methods. Robust data augmentation and class-balancing strategies ensured balanced performance across all lesion categories. Clinical validation by five board-certified Venezuelan dermatologists confirmed the system’s diagnostic accuracy, clinical relevance, and usability. The application’s deployment as an accessible web interface highlights its potential for supporting dermatological diagnosis, particularly in resource-limited settings. Our findings underscore the promise of deep learning-driven tools for enhancing skin cancer detection and improving healthcare equity in Latin America and beyond.

## 1 Introduction

Skin cancer represents one of the most prevalent malignancies worldwide, with melanoma accounting for the majority of skin cancer-related deaths despite being less common than other forms [1]. According to the World Health Organization [1], cancer is the leading cause of death globally, emphasizing the critical need for early detection tools to enable timely and appropriate care. The five-year survival rate for melanoma decreases significantly when detected in advanced stages, underscoring the importance of early diagnosis [2].

Dermatologists traditionally rely on visual inspection using dermoscopy—a non-invasive diagnostic technique that provides magnified, high-resolution images of skin lesions to evaluate their structural characteristics [3]. However, melanoma diagnosis remains subjective, imprecise, and can be extremely challenging for dermatologists without specialized training [4]. The interpretation of dermoscopic images by humans can be affected by incomplete visual search patterns, fatigue, distractions, and even the physical quality of the images [5, 6].

Recent advances in artificial intelligence (AI), particularly deep learning and convolutional neural networks (CNNs), have demonstrated remarkable success in medical image analysis. Esteva et al. [6] revealed that AI-based systems can match or even surpass expert dermatologists in detecting skin cancer. Similarly, in other medical imaging domains such as diabetic retinopathy and histopathology, deep learning algorithms have achieved diagnostic accuracy comparable to or exceeding that of trained specialists [7, 8].

The integration of medical imaging with computational methods has created a paradigm shift in healthcare, both in clinical practice and research. Digital processing of medical information has opened numerous possibilities for improving clinical diagnosis, making the use of appropriate tools for data capture by the medical community fundamental to leveraging the potential of image processing technologies applied in the medical field [9].

In dermatology, CNNs have shown significant growth in medical applications, particularly in the early detection and localization of dermatological diseases, overcoming the limitations of subjective human interpretation of images [10]. These AI-driven approaches offer efficient, non-invasive, computer-assisted diagnosis available continuously through accessible web interfaces, providing specialists with valuable decision-support tools for accurate diagnoses [5].

This technological advancement is particularly relevant in Venezuela and other Latin American countries, where healthcare systems face significant challenges, including limited access to specialized diagnostic technologies and shortages of trained dermatologists [11]. Specialists such as Dr. Margarita Oliver Llull and her colleagues at El Avila Clinic face limitations in accessing advanced technologies, making the implementation of AI-based diagnostic solutions urgent to enable more accurate and effective diagnoses. A support system of these characteristics would not only contribute to improving diagnostic quality but also optimize response times and treatment outcomes, positively impacting patient care [11].

Building upon this foundation, the present study aims to develop and clinically validate an intelligent system capable of detecting and classifying nine types of cutaneous lesions, including melanoma, actinic keratosis, basal cell carcinoma, dermatofibroma, nevus, benign pigmented keratosis, seborrheic keratosis, squamous cell carcinoma, and vascular lesions using deep learning based on dermoscopic image processing. The system integrates state-of-the-art CNN architectures (NFNet-L0, ResNeSt-101e, and MogaNet-XT) and provides an accessible web interface for clinical use, validated by board-certified Venezuelan dermatologists.

Unlike previous studies focusing on binary classification or limited lesion types, this work addresses a multiclass classification task over nine distinct dermatological conditions, offering a comprehensive diagnostic tool. Moreover, clinical validation in a resource-limited setting provides unique insights into the practical applicability of AI-assisted diagnosis in underserved regions. The deployment of the system as an open-access web application on Hugging Face ensures accessibility and democratizes advanced diagnostic capabilities worldwide.

## 2 Related Works

Automated diagnosis of skin cancer using machine learning and deep learning techniques has been extensively studied in recent years. Traditional manual diagnosis through dermoscopy has inherent limitations including subjective interpretation and reliance on specialist expertise [3, 4].

Several studies have demonstrated the capability of convolutional neural networks (CNNs) to classify various types of skin lesions with high accuracy. Esteva et al. [6] pioneered the use of deep CNNs for skin cancer classification, showing that AI systems can reach dermatologist-level performance. Subsequent works such as Gulshan et al. [7] and Bejnordi et al. [8] confirmed the efficacy of deep learning in medical image analysis across different domains, reinforcing its applicability to dermatology.

More recent advances include the use of novel and hybrid CNN architectures to improve classification performance on imbalanced and diverse datasets. Networks such as NFNet, ResNeSt, and MogaNet have demonstrated robustness in feature extraction and classification power in medical imaging applications [12]. Specifically in melanoma and cutaneous lesion diagnosis, multiple studies have emphasized the importance of using large, well-annotated datasets combining public archives and clinical images [13].

Despite the promising results, few studies have integrated clinical validation with dermatologists in resource-limited settings, a gap this work addresses by involving experts from El Avila Clinic and embedding the system in an accessible web platform for real-time use.

These advancements underline the potential of AI-assisted diagnosis to enhance accuracy and accessibility in dermatological care, motivating the development of the system presented in this study.

## 3 Methodology

### 3.1 Dataset Description

The dataset employed in this study was assembled from two primary sources: the publicly available International Skin Imaging Collaboration (ISIC) archive and clinical images obtained from patients at El Avila Clinic in Caracas, Venezuela, provided by Dr. Margarita Oliver Llull. The initial dataset comprised 2,357 dermoscopic images encompassing nine distinct lesion classes: melanoma, actinic keratosis, basal cell carcinoma, dermatofibroma, melanocytic nevus, benign pigmented keratosis, seborrheic keratosis, squamous cell carcinoma, and vascular lesions.

The ISIC archive is a well-established repository in dermatological research, known for its extensive collection of high-quality images accompanied by verified clinical annotations. The images sourced from ISIC contributed significantly to the diversity of the dataset in terms of skin tones, lesion types, and acquisition conditions. Clinical images contributed by El Avila Clinic were collected under approved ethical protocols, ensuring patient confidentiality and adherence to data privacy regulations. All clinical images underwent a manual annotation process by experienced dermatologists, guided by consensus criteria, to verify lesion types, thus providing reliable ground truth labels for the supervised learning task.

During the data preparation phase, 133 corrupted images were identified and removed to ensure dataset integrity and quality. The resulting dataset exhibited significant class imbalance, with certain lesion classes substantially underrepresented compared to others (see Figure 2), posing a challenge for supervised learning models that can become biased towards majority classes.

Sample images illustrating representative lesions from each class are shown in Figure 1, highlighting the visual variability and complexity characterizing the classification challenge.

**Figure 1.**
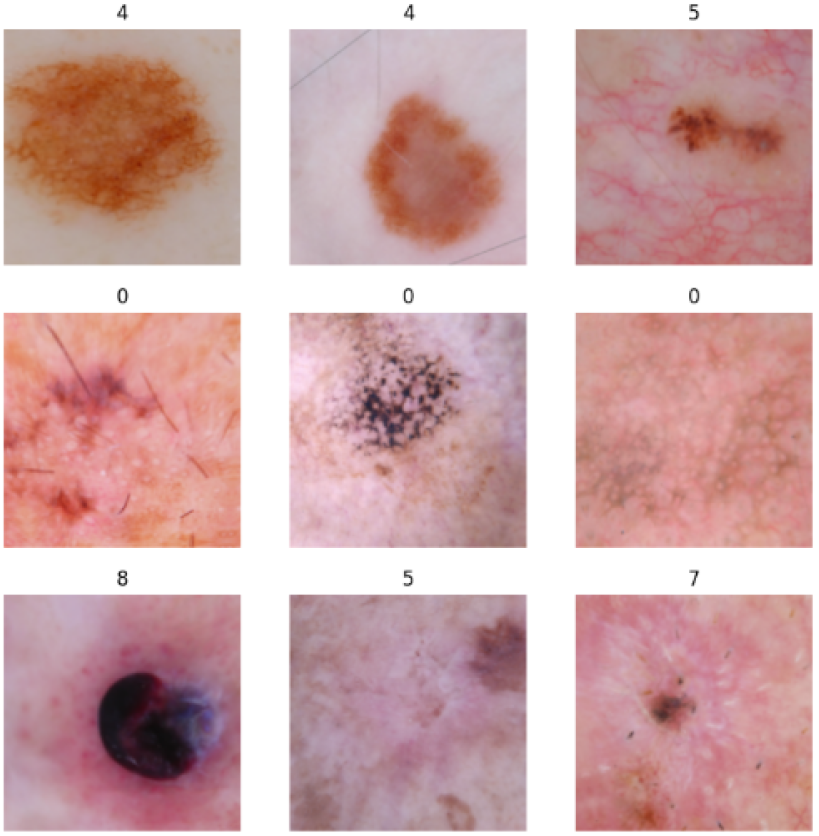
Representative dermoscopic images from each lesion category illustrating the visual diversity and complexity.

**Figure 2.**
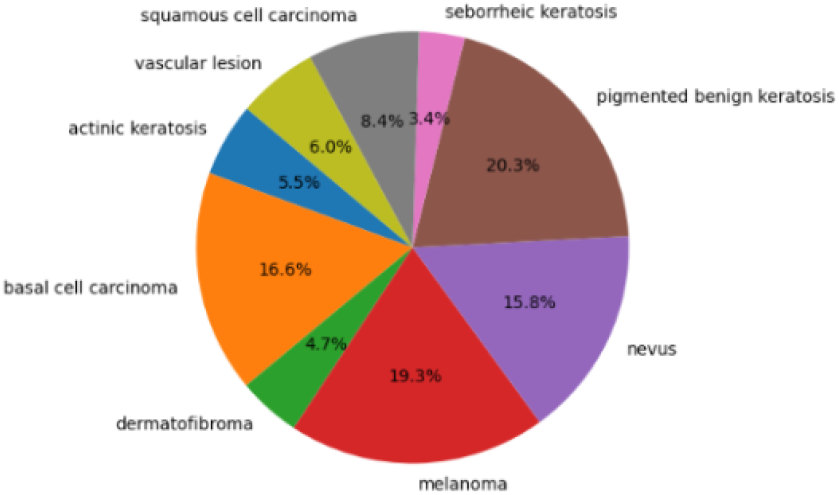
Graph displaying the class imbalance present in the original dataset prior to preprocessing and augmentation.

This diversified dataset provided the foundation for subsequent preprocessing and augmentation to create a balanced training corpus for convolutional neural network architectures.

### 3.2 Data Preprocessing and Augmentation

The original dataset of 2,357 dermoscopic images exhibited significant class imbalance, with certain lesion types vastly outnumbering others. Figure 2 illustrates the severe imbalance in the distribution of lesion classes during the exploratory data analysis. This imbalance poses a challenge for supervised learning models, as models can become biased towards majority classes, leading to poor generalization for underrepresented lesions.

To mitigate this issue, an extensive data preprocessing and augmentation pipeline was implemented. Initially, all images were resized to a uniform 224 x 224-pixel dimension using bicubic interpolation to conform with the CNN input requirements while preserving critical spatial features. Pixel intensity normalization was performed to rescale pixel values to a [0,1] range, reducing the impact of varying lighting conditions and acquisition artifacts while expediting model training convergence.

To address class imbalance, a data augmentation strategy was employed to balance the dataset. A minimum threshold of 2,500 images per class was established, and synthetic images were generated for underrepresented classes through random sampling and augmentation of existing samples. Augmentation transformations included random flips (horizontal and vertical), rotations within ±30 degrees, zooming with scale variations between 0.8 and 1.2, brightness and contrast adjustments, and addition of Gaussian noise. These transformations simulate diverse imaging conditions and lesion presentations, encouraging model robustness.

The augmentation process significantly enhanced dataset balance, as depicted in Figure 3 and Table 1, resulting in a final balanced dataset of 22,618 images distributed uniformly across the nine lesion classes.

**Table 1.** Distribution of dermoscopic images per lesion category after augmentation.

| Lesion | Images |
| --- | --- |
| Melanoma | 2516 |
| Actinic Keratosis | 2516 |
| Basal Cell Carcinoma | 2516 |
| Dermatofibroma | 2516 |
| Melanocytic Nevus | 2516 |
| Benign Pigmented Keratosis | 2516 |
| Seborrheic Keratosis | 2503 |
| Squamous Cell Carcinoma | 2516 |
| Vascular Lesions | 2503 |
| Total | 22618 |

**Figure 3.**
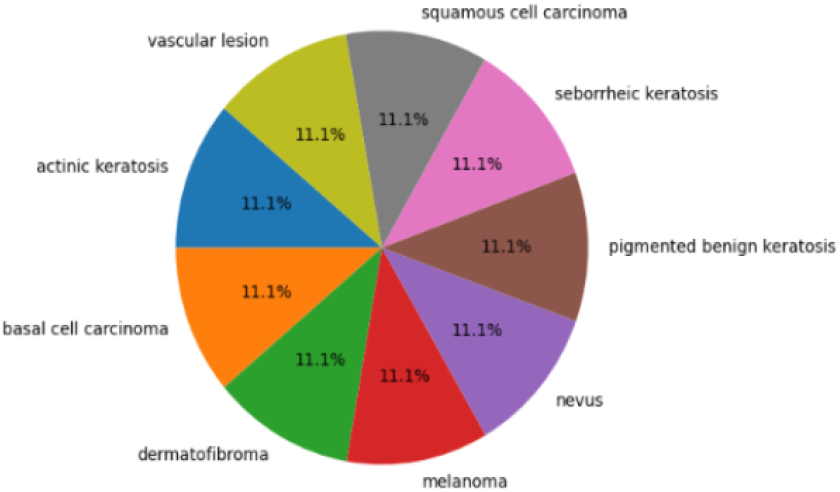
Graph showing the balanced distribution of classes achieved after preprocessing and augmentation.

In parallel, batch sampling during training incorporated oversampling of minority classes and undersampling of majority classes to maintain balanced class representation in each minibatch. This strategy prevented model over-fitting to dominant classes and facilitated more equitable learning across all lesion categories.

Additional preprocessing steps included sharpening of images using a Laplacian filter to enhance lesion edge boundaries, and contrast-limited adaptive histogram equalization (CLAHE) to improve local contrast and highlight subtle lesion features. These enhancements aided the CNNs’ capability to extract meaningful features for accurate classification.

Collectively, these preprocessing and augmentation methodologies addressed the challenges of dataset diversity, class imbalance, and imaging variability, resulting in a high-quality and balanced dataset that maximized model learning potential and generalization across nine distinct cutaneous lesion categories.

### 3.3 CNN Architectures Used

Convolutional Neural Networks (CNNs) constitute the foundational architecture employed in this research for automated melanoma and cutaneous lesion classification. CNNs are designed to automatically and adaptively learn spatial hierarchies of features through stacked convolutional layers, activation functions, and pooling mechanisms that mimic the hierarchical processing of the human visual cortex.

In this work, three advanced CNN architectures were implemented and evaluated: NFNet-L0, ResNeSt-101e, and MogaNet-XT. These were chosen based on their recent state-of-the-art performance on image classification benchmarks and their complementary architectural innovations.

#### 3.3.1 NFNet-L0

NFNet (Normalization-Free Network), proposed by Brock et al. [12], introduces a novel training method that removes the need for batch normalization through adaptive gradient clipping, allowing for stable training with large batch sizes. This culminates in improved robustness and faster convergence. The NFNet-L0 variant represents a high-capacity model balancing speed and accuracy.

Figure 4 presents a detailed view of the NFNet transition block, illustrating the complex residual connections and adaptive gain scaling employed.

**Figure 4.**
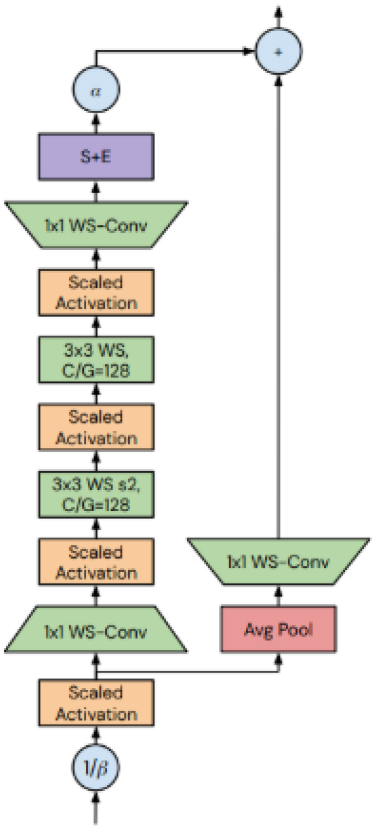
NFNet Transition Block Architecture

The adaptive gradient clipping leverages the unit-wise ratio of gradient norms to parameter norms to maintain stable updates without the normalization layers [12]. Mathematically, this controlled clipping is expressed as:

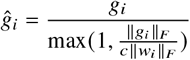

where *g*_*i*_ is the gradient for unit *i, w*_*i*_ are the corresponding parameters, ∥ · ∥ _*F*_ denotes the Frobenius norm, and *c* is the clipping coefficient (typically set to 0.01).

#### 3.3.2 ResNeSt-101e

ResNeSt (Split-Attention Network), developed by Zhang et al. [14], enhances the ResNet architecture by incorporating a split-attention mechanism that captures cross-channel feature interactions through multiple cardinal groups.

The detailed split-attention unit is illustrated in Figure 5. This unit allows the network to weigh features across cardinal splits adaptively, improving representational power.

**Figure 5.**
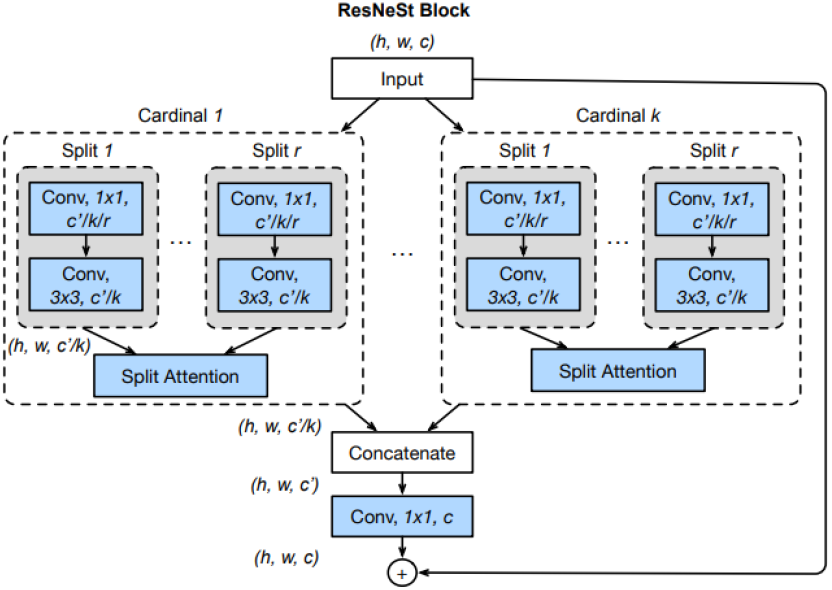
Split-Attention Module of ResNeSt

The split-attention operation for the *c*-th channel of the *k*-th cardinal group is formalized as:

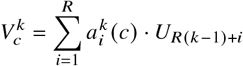

where *U*_*R*(*k* − 1) +*i*_ represents the feature-map splits, *R* is the radix (number of splits), and 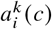 are the learned attention weights computed via softmax over the splits [14].

#### 3.3.3 MogaNet-XT

MogaNet, introduced by Li et al. [15], is a multi-order gated aggregation CNN focusing on efficient extraction of contextual information by combining spatial and channel aggregation blocks.

Figure 6 depicts the general structure combining spatial and channel aggregations.

**Figure 6.**
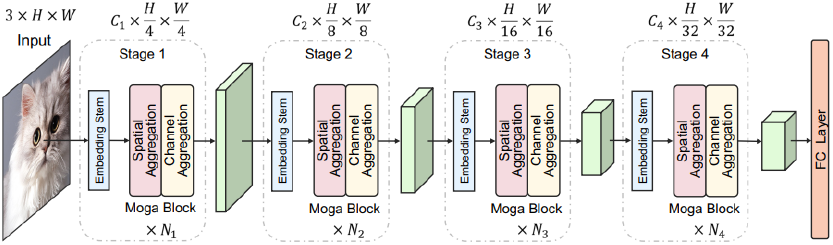
Spatial and Channel Aggregation Blocks in MogaNet

The model employs multi-order gated aggregation where features from spatial and channel dimensions are adaptively fused. The spatial aggregation block processes multi-order spatial interactions, while the channel aggregation module reallocates channel-wise features. The combined output is represented by:

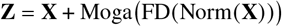

where FD(·) is a feature decomposition module, Moga(·) denotes the multi-order gated aggregation comprising gating and context branches, and **X** is the input feature [15].

#### 3.3.4 Summary

These three architectures provide complementary strengths: NFNet-L0 brings normalization-free training stability; ResNeSt employs attention-based feature weighting for enhanced representation; and MogaNet leverages multi-order gated aggregation for contextual awareness.

Our experiments utilize these models trained with the same dataset and optimization protocol to evaluate their comparative classification performance on melanoma and other cutaneous lesions.

### 3.4 Training Procedure

The dataset was split into training, validation, and test sets using an 80%/10%/10% stratified sampling strategy to maintain class balance across partitions. This split ensured that the models were trained and validated on representative subsets while preserving unseen data for final evaluation.

Training leveraged the PyTorch deep learning framework [16] with the timm library [17] to facilitate model implementations and transfer learning capabilities. The base model for the NFNet-L0 architecture was pretrained on the ImageNet-1k dataset, consisting of over 1.28 million images spanning 1,000 classes [18]. Transfer learning facilitated leveraging pretrained feature extractors, enabling faster convergence and improved performance on the melanoma dataset.

A custom classification head was appended to each base model. This head consisted of an adaptive average pooling layer transforming convolutional feature maps to a fixed size, followed by a flattening operation and a fully connected linear layer mapping to the nine lesion classes.

To optimize the models, the cross-entropy loss function was employed as the loss criterion, widely used in multi-class classification tasks to measure the dissimilarity between predicted and actual class probabilities [19]. The Adam optimizer was selected for its adaptive learning rates, accelerating convergence while maintaining training stability [20].

An optimal learning rate was identified via an exploratory learning rate range test [21] (Figure 7), ensuring efficient convergence during training with a balanced tradeoff between speed and stability.

**Figure 7.**
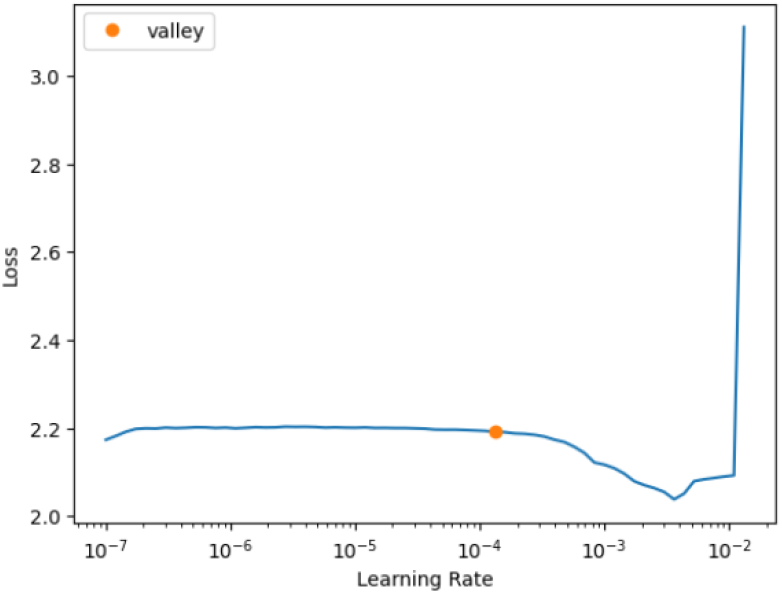
Learning rate finder plot used to determine optimal learning rate.

To reduce overfitting while harnessing transfer learning benefits, the base pretrained layers were frozen for the first several epochs, training only the newly added classification head. Subsequently, all layers were unfrozen for fine-tuning, allowing the entire network to adapt to specific features of the skin lesion dataset. This progressive unfreezing strategy improved training efficiency and accuracy, especially on the relatively small labeled dataset.

Training was conducted on an NVIDIA RTX 2080 SUPER GPU environment, with each epoch lasting approximately 28 minutes for the NFNet-L0 model. The training spanned 17 epochs in total, with early stopping based on validation loss.

Due to resource constraints, training of the ResNeSt-101e model was limited to 6 epochs. Consequently, comparative evaluations primarily focused on the NFNet-L0 and MogaNet-XT models, which were fully trained for 17 epochs.

Tables 2, 3, and 4 present detailed training results for NFNet-L0, ResNeSt-101e, and MogaNet-XT, respectively. The NFNet-L0 model achieved superior performance, reaching a final accuracy of 96.17%, supported by a high F1-score and precision, indicating strong predictive capability (see Table 2).

**Table 2.** Training results for NFNet-L0 across iterations.

| Iter. | Train Loss | Val Loss | Accuracy |
| --- | --- | --- | --- |
| 0 | 1.936 | 1.900 | 0.366 |
| 1 | 1.129 | 1.215 | 0.551 |
| 2 | 0.847 | 0.892 | 0.656 |
| 16 | 0.110 | 0.124 | 0.962 |

**Table 3.** Training results for ResNeSt-101e.

| Iter. | Train Loss | Val Loss | Accuracy |
| --- | --- | --- | --- |
| 0 | 0.995 | 1.935 | 0.415 |
| 1 | 0.550 | 45.795 | 0.462 |
| 2 | 0.487 | 0.834 | 0.708 |
| 5 | 0.137 | 0.210 | 0.923 |

**Table 4.**
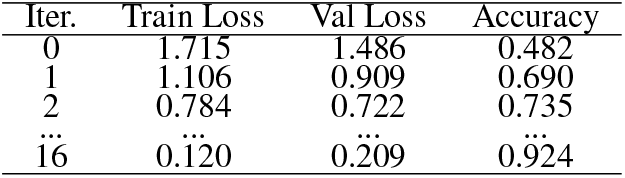
Training results for MogaNet-XT.

Finally, Table 5 compares the performance between the NFNet-L0 and MogaNet-XT models after full training. NFNet-L0 demonstrated superior accuracy and generalization capabilities, supporting its use as the optimal model for clinical deployment.

**Table 5.** Comparative performance of NFNet-L0 and MogaNet-XT.

| Model | Final Loss | Accuracy |
| --- | --- | --- |
| MogaNet-XT | 0.121 | 0.924 |
| NFNet-L0 | 0.103 | 0.962 |

**Table 6.** Detailed evaluation metrics and confusion matrix for NFNet-L0 on the test set.

| Metric | Value | Description |
| --- | --- | --- |
| Accuracy | 0.962 | Overall correctness of predictions |
| Precision | 0.967 | Proportion of positive identifications that were correct |
| Recall (Sensitivity) | 0.960 | Proportion of actual positives identified correctly |
| F1 Score | 0.961 | Harmonic mean of precision and recall |
| Specificity | 1.0 | Proportion of negatives correctly identified |

|  | MEL | AK | BCC | DF | NV | BPK | SK | SCC | VASC |
| --- | --- | --- | --- | --- | --- | --- | --- | --- | --- |
| MEL | 443 | 0 | 0 | 3 | 0 | 0 | 0 | 0 | 0 |
| AK | 0 | 468 | 0 | 0 | 0 | 0 | 1 | 0 | 0 |
| BCC | 0 | 0 | 477 | 0 | 0 | 0 | 0 | 1 | 0 |
| DF | 0 | 0 | 0 | 335 | 8 | 2 | 65 | 1 | 0 |
| NV | 75 | 0 | 0 | 0 | 383 | 0 | 0 | 0 | 0 |
| BPK | 0 | 0 | 0 | 0 | 0 | 426 | 0 | 0 | 0 |
| SK | 0 | 0 | 0 | 0 | 0 | 0 | 498 | 0 | 0 |
| SCC | 0 | 0 | 0 | 0 | 1 | 0 | 1 | 452 | 0 |
| VASC | 0 | 0 | 0 | 0 | 0 | 0 | 0 | 0 | 437 |

The NFNet-L0 model demonstrated exceptional performance across all measured metrics, highlighting its suitability as the primary model for clinical deployment.

These comprehensive results provide a solid foundation for evaluating and comparing future model iterations.

### 3.5 Evaluation Metrics

Model performance was assessed using a comprehensive set of classification metrics, following the methodologies of Tharwat [22] and Villanueva [23]. These metrics provide a robust framework for evaluating and comparing the effectiveness of deep learning classifiers in medical imaging tasks.

#### Confusion Matrix

The confusion matrix is a fundamental tool that summarizes model predictions with respect to actual classes, condensing the multi-class classification outcome into a visual summary. Each row represents the true class, while each column represents the predicted class (see Figure 8, adapted from Tharwat [22]).

**Figure 8.**
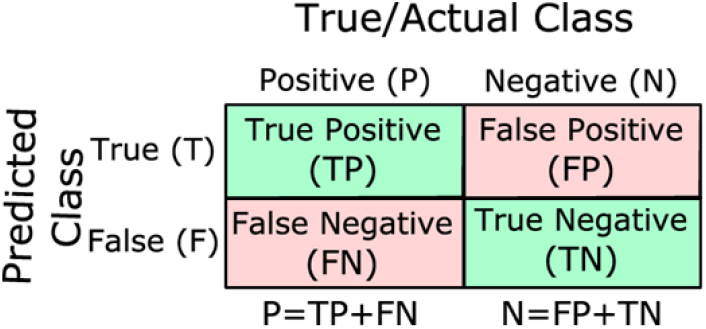
Architecture of a confusion matrix adapted from Tharwat [22].

From the confusion matrix, four main counts are derived:

- True Positive (TP): Correct positive predictions.
- True Negative (TN): Correct negative predictions.
- False Positive (FP): Incorrect positive predictions (predicted positive but actually negative).
- False Negative (FN): Incorrect negative predictions (predicted negative but actually positive).

**Accuracy (ACC)** is the proportion of correctly classified samples over the total number of samples visualized:

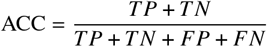

**Precision (PREC)** is the proportion of positive identifications that were actually correct:

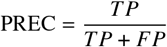

**Sensitivity (SEN) / Recall** is the proportion of actual positives correctly identified:

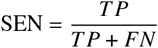

**Specificity (SPC)** is the proportion of negatives correctly identified:

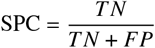

**F1 Score (F-measure)** is the harmonic mean of precision and sensitivity, providing an overall measure of a model’s balance between precision and recall:

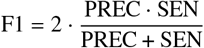

These performance metrics were computed on the independent test set after final model training. The combination of accuracy, precision, sensitivity, specificity, and F1 score facilitates a holistic understanding of the clinical and statistical significance of the deep learning classification results.

The interpretations and formulas above follow the protocols described in Tharwat [22] for confusion matrix-based performance, and Villanueva [23] for clinical model evaluation in melanoma studies.

### 3.6 Clinical Validation Protocol

Clinical validation of the artificial intelligence model was undertaken in collaboration with five board-certified Venezuelan dermatologists, coordinated through Dr. Margarita Oliver Llull at El Avila Clinic. An initial cohort of ten dermatologists from leading clinics in Caracas was contacted; ultimately five experts participated within the available four-week evaluation period.

Validation employed a structured survey distributed via email, accompanied by dermatological image sets processed by the deep learning model. Participants were asked to review the model’s predictions and complete the survey, assessing multiple aspects including diagnostic accuracy, clinical utility of the tool, reliability of outputs, and usability of the interface. The evaluation process was designed to reflect real clinical scenarios and decision-making contexts.

Figure 9 demonstrates the overall model precision as assessed by the participating dermatologists.

**Figure 9.**
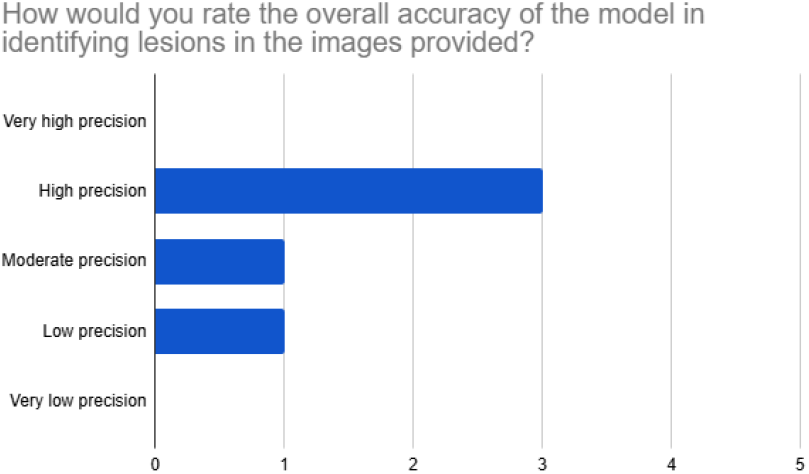
Overall precision of the model according to clinical evaluation by dermatologists.

Clinical utility for diagnostic support was visualized (see Figure 10), while relevance and reliability of the model’s recommendations were summarized in Figure 11. Interpretability and ease of use were highly rated by experts, with the majority reporting that model predictions were clear and the user interface was intuitive (Figure 12; Figure 13).

**Figure 10.**
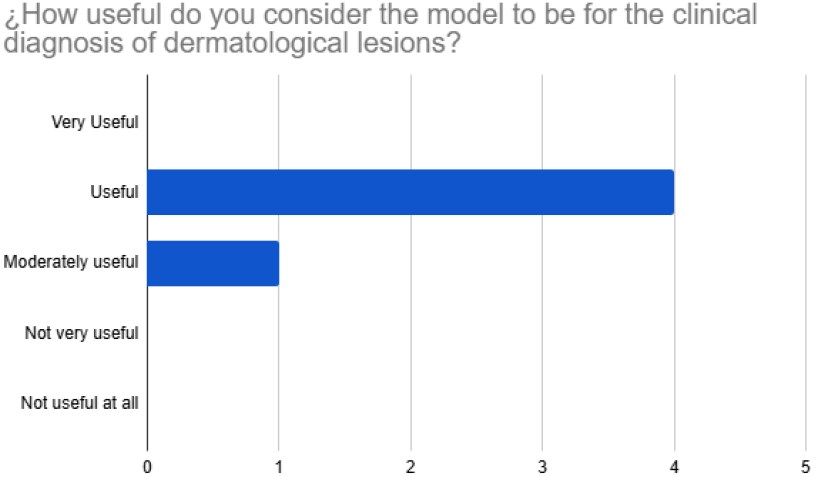
Clinical utility of the model for dermatological diagnosis.

**Figure 11.**
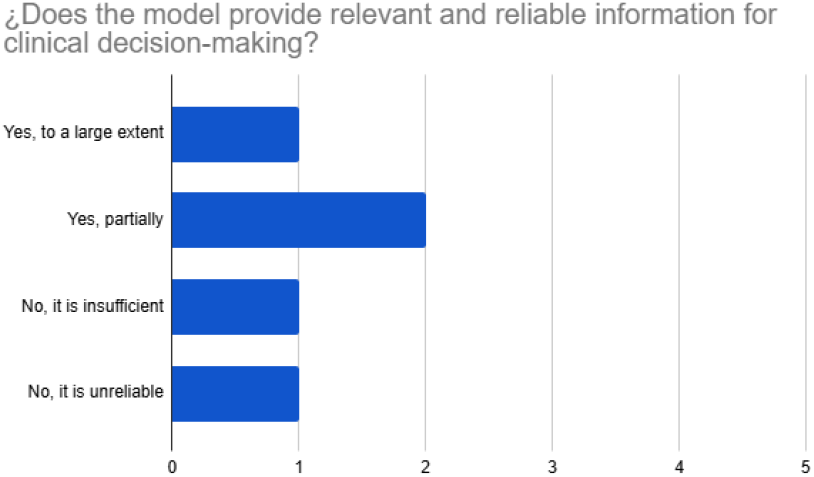
Relevance and reliability of information provided by the model.

**Figure 12.**
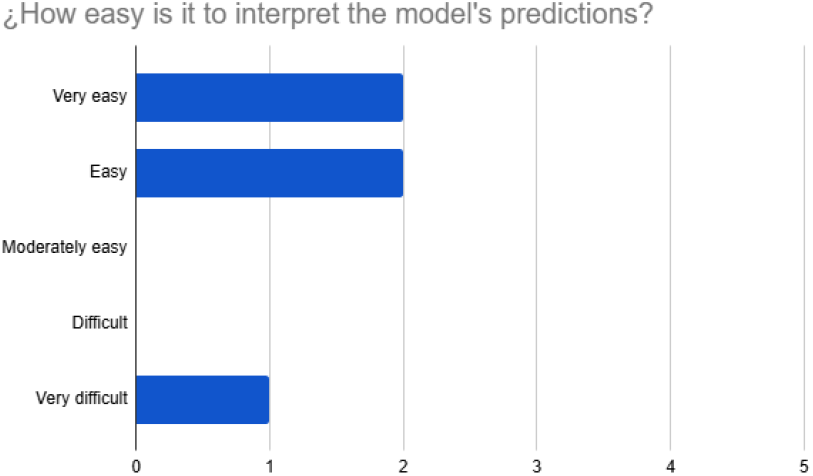
Ease of interpretation of model predictions as rated by dermatologists.

**Figure 13.**
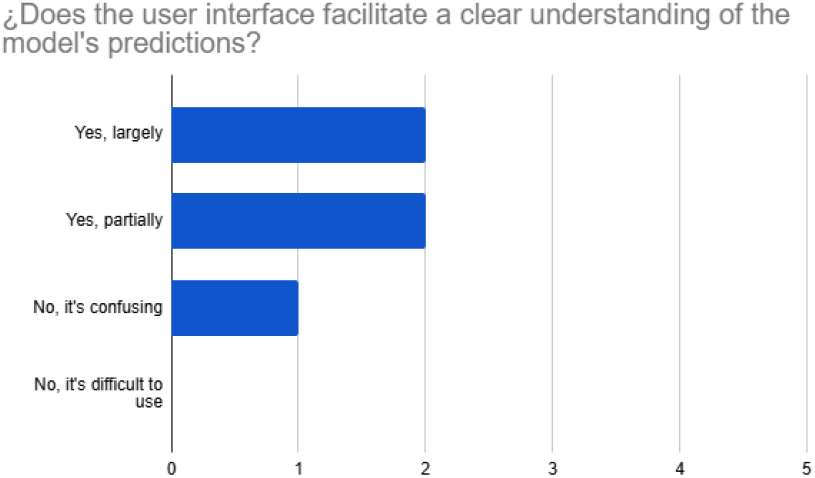
Efficiency of the user interface for understanding model outputs.

Feedback also included constructive recommendations for future system improvement, such as allowing multi-image inputs during inference, expanding the lesion database, and offering offline access to accommodate limited internet connectivity in clinical practice.

In summary, clinical evaluations highlighted high diagnostic accuracy and clinical usefulness of the AI tool, with dermatologists affirming its value for supporting dermatological decision-making. Recommendations from clinical feedback present actionable next steps for refining and deploying the system in broader, resource-variable settings.

## 4 Conclusion

This study presents a robust, clinically validated deep learning system for the automated classification of melanoma and eight additional cutaneous lesions, leveraging state-of-the-art convolutional architectures such as NFNet-L0, ResNeSt-101e, and MogaNet-XT. Through rigorous preprocessing, targeted augmentation, and transfer learning, our NFNet-L0-based model achieved a benchmark accuracy of 96.2%, demonstrating outstanding reliability across diverse skin lesion categories and patient populations.

Clinical validation by Venezuelan dermatologists at El Avila Clinic confirmed the system’s diagnostic accuracy, practical utility, and interpretability in real-world medical settings. The collaborative survey and usability study highlighted the model’s readiness for clinical deployment as a decision support tool, particularly in resource-constrained environments across Latin America.

Our results reinforce the critical potential of AI-driven diagnostic tools to augment dermatologist workflows, bridge gaps in healthcare access, and empower early detection of skin cancer. Importantly, the model is deployed as an open-access web application on the Hugging Face Spaces platform (https://huggingface.co/spaces/dcavadia/MelanoScopeAI), enhancing accessibility for clinicians and researchers worldwide.

Future work will focus on expanding the dataset, enhancing multi-image inference capabilities, and incorporating offline usability for greater clinical reach. The constructive feedback from practicing experts will continue to shape model improvements, promoting safe, equitable, and effective AI adoption in global dermatology.

In summary, our clinically validated AI model sets a new standard for deep learning-enabled skin cancer screening, offering significant contributions to the field of medical AI and demonstrating tangible benefits for patients and practitioners worldwide.

## Author Contributions

Daniel Cavadia designed and implemented the AI model, conducted experiments, and wrote the manuscript. Marcel Castro provided academic supervision and methodological guidance. Margarita Oliver Llull facilitated clinical data acquisition and supervised clinical validation.

## Ethics Statement

This study utilized dermoscopic images from two sources: the publicly available International Skin Imaging Collaboration (ISIC) archive and clinical images provided by Dr. Margarita Oliver Llull from El Avila Clinic, Caracas, Venezuela. All clinical images were fully anonymized prior to analysis in accordance with institutional ethical guidelines, and informed consent for research use was obtained from patients. Participation of dermatologists in the validation survey was voluntary, and written consent was obtained from all participants. This research was conducted in accordance with the Declaration of Helsinki and ethical guidelines for human subjects research.

## Data Availability Statement

The ISIC dermoscopic image dataset used in this study is publicly available at https://www.isic-archive.com. Clinical images from El Avila Clinic, Caracas, Venezuela, are not publicly available due to patient privacy and institutional data sharing restrictions but may be available from the corresponding author upon reasonable request and with appropriate ethical approval.

